# Diagnostic information from a negative QuantiFERON-TB Gold Plus result in adults evaluated for tuberculosis in Brazil and South Africa

**DOI:** 10.64898/2026.09.25.26363952

**Authors:** Allyson Guimarães Costa, Anete Trajman, Maria Cecília Borges-Cabral, Aliasgar Esmail, Brenda Karoline Souza Carvalho, Bruna Pires Loiola, Alexandra Brito Souza, Renata Spener-Gomes, Jaquelane Silva Jesus, Vanderson Souza Sampaio, Daniel Barros de Castro, Keertan Dheda, Marcelo Cordeiro-Santos

## Abstract

Interferon-gamma release assays detect *Mycobacterium tuberculosis* infection but cannot distinguish infection from active tuberculosis (TB). We evaluated the diagnostic information provided by QuantiFERON-TB Gold Plus (QFT-Plus) among adults investigated for active TB in Brazil and South Africa. Adults with presumptive TB were prospectively enrolled by convenience sampling. QFT-Plus was the index test, and diagnostic classification required microbiological confirmation of TB or an established alternative diagnosis. QFT-Plus results did not inform diagnostic classification. Accuracy was assessed among participants with determinate results, primarily using likelihood ratios. Of 723 participants tested, 48 (6.6%) had indeterminate results, all due to low mitogen responses. Among 675 participants with determinate results, 147 (21.8%) had confirmed TB. Sensitivity was 68.0% (95% CI, 60.1–75.0), specificity was 59.3% (95% CI, 55.0–63.4), and the negative likelihood ratio was 0.54 (95% CI, 0.42– 0.69). At the observed prevalence, a negative result left a 13.1% probability of active TB. Negative likelihood ratio confidence intervals included 1 in both South African HIV strata. Specificity was higher among participants with HIV in both countries. Adding TB2 to TB1 alone identified 15 additional TB cases and generated 18 additional positive results among participants without active TB. HIV and active TB were independently associated with indeterminate results in exploratory regression. A negative QFT-Plus result provided insufficient information to exclude active TB. Higher negative predictive values among participants with HIV reflected lower disease prevalence and higher specificity rather than greater sensitivity. Negative or indeterminate results should not terminate diagnostic evaluation.

**IMPORTANCE:** Blood tests that detect immune responses to *Mycobacterium tuberculosis* can yield negative results in people with active tuberculosis. Understanding how much reassurance a negative result provides is therefore important when these tests are encountered during diagnostic investigation. Among symptomatic adults in Brazil and South Africa, a negative QuantiFERON-TB Gold Plus result left a substantial probability of active disease. Differences between populations showed why apparently reassuring negative predictive values can be misleading. The additional antigen tube detected more cases but did not overcome the test’s limited ability to exclude disease. Indeterminate results were associated with human immunodeficiency virus infection and active tuberculosis. These findings support laboratory reporting and clinical interpretation that clearly communicate the limitations of negative and indeterminate results and the need for continued investigation when active tuberculosis remains suspected.

## INTRODUCTION

Tuberculosis (TB) remains a leading cause of illness and death worldwide. In 2024, an estimated 10.7 million people developed TB and 1.23 million died from the disease, including approximately 150,000 deaths among people with HIV. People living with HIV were approximately 12 times more likely to develop TB than those without HIV, with the greatest burden of HIV-associated TB occurring in the African Region [1]. Diagnosis can be particularly challenging in this population because immunosuppression may be associated with paucibacillary pulmonary disease, extrapulmonary involvement, atypical presentations, and reduced sensitivity of microbiological tests [2]. A non-sputum-based test providing sufficient information to exclude active TB could therefore contribute to the diagnostic evaluation of adults with presumptive TB.

Interferon-gamma release assays (IGRAs) measure cell-mediated immune responses to *Mycobacterium tuberculosis*-specific antigens and were developed to identify TB infection as an alternative to the tuberculin skin test. Because IGRAs cannot distinguish TB infection from active disease, they are not recommended for diagnosing or excluding active TB [2,3]. QuantiFERON-TB Gold Plus (QFT-Plus) contains two antigen tubes. TB1 is designed primarily to elicit CD4^+^ T-cell responses, whereas TB2 contains additional shorter peptides intended to elicit responses from both CD4^+^ and CD8^+^ T cells [4,5]. Nevertheless, the difference between TB2 and TB1 is not a direct measure of CD8⁺ T-cell activity, and the clinical contribution of TB2 remains uncertain [4].

Studies of QFT-Plus in patients with active TB have reported heterogeneous sensitivity. An earlier meta-analysis estimated a pooled sensitivity of approximately 94% but found no significant improvement over the previous QuantiFERON-TB Gold In-Tube assay [6]. A subsequent meta-analysis similarly found no clear diagnostic-performance advantage over QuantiFERON-TB Gold In-Tube or T-SPOT.TB [7]. Studies in high-burden settings have demonstrated substantially lower sensitivity, with negative IGRA results occurring in patients with microbiologically confirmed TB [3,8]. HIV-related immunosuppression may further impair antigen and mitogen responses, increasing the frequency of false-negative or indeterminate results [9]. Evidence concerning the incremental contribution of TB2 is also inconsistent, and its magnitude and clinical relevance appear to vary across populations [4,10].

Although QFT-Plus was developed to detect M. tuberculosis infection, its results may be available during investigation for active TB. Understanding how much a negative result changes disease probability, and how this varies across clinical populations, is therefore relevant to test interpretation. Likelihood ratios quantify the change from pretest to post-test odds and, unlike predictive values, are not mathematically determined by disease prevalence. However, they may vary with the clinical characteristics of the population tested. Thus, we evaluated the diagnostic information provided by QFT-Plus among adults with presumptive TB in Brazil and South Africa, focusing on the negative likelihood ratio, residual post-test probability, and variation by country and HIV status. Secondary analyses examined the incremental contribution of TB2, quantitative TB1 and TB2 responses, and the frequency and correlates of indeterminate results.

## MATERIALS AND METHODS

### Study design and setting

We conducted a prospective, multicentre, cross-sectional diagnostic-accuracy analysis embedded within ImmiPrint-TB, a larger study evaluating a blood-based host-response biomarker signature for distinguishing active TB from alternative diagnoses among symptomatic adults. Participants were recruited in Manaus, Brazil, and Cape Town, South Africa, from May to December 2018. In Manaus, recruitment occurred at Fundação de Medicina Tropical Doutor Heitor Vieira Dourado, a referral centre for tropical diseases and HIV care, and Fundação de Pneumologia Sanitária Cardoso Fontes, a referral centre for TB and respiratory diseases. The former predominantly served people with HIV, whereas the latter received patients referred for TB investigation. In Cape Town, participants were recruited at four primary healthcare facilities.

QFT-Plus was performed as part of ImmiPrint-TB because one of the host-response biomarker analyses used plasma from the TB1 and TB2 antigen-stimulated tubes. In the present analysis, QFT-Plus was evaluated separately as the index test, and the final diagnostic classification established within the parent study served as the reference standard. QFT-Plus results were unavailable to clinicians and did not influence diagnostic or therapeutic decisions. Molecular testing was performed using Xpert MTB/RIF in both countries according to the predefined study procedures.

### Ethical considerations

The Brazilian component was approved by the Research Ethics Committee of Fundação de Medicina Tropical Doutor Heitor Vieira Dourado (approval no. 2,525,182; CAAE 80643917.4.0000.5260), and the South African component by the University of Cape Town Human Research Ethics Committee (HREC reference 290/2020). All procedures complied with applicable national regulations and the Declaration of Helsinki. All participants provided written informed consent before enrolment.

### Participants and diagnostic classification

Adults aged ≥18 years with symptoms compatible with TB were enrolled in the parent ImmiPrint-TB study when study personnel were available, constituting an operational convenience sample. Participants underwent assessment for active TB and were offered rapid HIV testing. QFT-Plus sampling and microbiological investigation were performed during the same diagnostic episode. Final diagnostic classification was established after completion of the diagnostic investigation and clinical follow-up.

Eligibility for the present analysis required a QFT-Plus result and a valid final diagnostic classification. Participants lacking either requirement or in whom nontuberculous mycobacteria were identified were excluded. Active TB was defined by detection of *Mycobacterium tuberculosis* complex using Xpert MTB/RIF or mycobacterial culture in a respiratory or non-respiratory specimen. Participants without microbiological evidence of TB were classified as not having active TB when an alternative diagnosis was established through clinical assessment, laboratory and radiological findings, and clinical follow-up.

The analysis of indeterminate results included all eligible participants, whereas the primary diagnostic-accuracy analysis was restricted to those with determinate QFT-Plus results.

### Data collection

Trained study personnel collected sociodemographic and clinical data using a standardised questionnaire. Laboratory results related to TB investigation, CD4^+^ T-cell counts, and HIV viral loads were retrieved from study records and electronic medical records when available. Missing observations were excluded from the relevant analyses, resulting in variable denominators. Adverse events related to the index test and reference-standard procedures were not systematically assessed.

### Microbiological procedures

Clinical specimens were processed on the day of collection. When appropriate, specimens underwent digestion and decontamination using the N-acetyl-L-cysteine– sodium hydroxide method, following established mycobacteriology and biosafety procedures [11, 12]. Xpert MTB/RIF testing was performed on the GeneXpert system (Cepheid, Sunnyvale, CA, USA) according to the manufacturer’s instructions. For mycobacterial culture, specimens were inoculated into Mycobacteria Growth Indicator Tubes and incubated at 37°C in the automated BACTEC MGIT system (Becton Dickinson, Sparks, MD, USA). Positive cultures were evaluated to confirm *M. tuberculosis* complex.

### QFT-Plus procedures and interpretation

Venous blood was collected into Nil, TB1, TB2, and Mitogen tubes and incubated at 37°C for approximately 20 h. Plasma was then separated and stored at −20°C until testing. Interferon-gamma concentrations were measured in IU/mL using the QFT-Plus enzyme-linked immunosorbent assay, and results were interpreted according to the manufacturer’s instructions [5].

Background-corrected responses were calculated as TB1−Nil, TB2−Nil, and Mitogen−Nil. Results were positive when Nil was ≤8.0 IU/mL and at least one antigen tube had a background-corrected response ≥0.35 IU/mL and ≥25% of the Nil value. Results were negative when neither antigen tube met the positivity criteria, Nil was ≤8.0 IU/mL, and Mitogen−Nil was ≥0.50 IU/mL. Results were indeterminate when Nil was >8.0 IU/mL or when neither antigen tube met the positivity criteria and Mitogen−Nil was <0.50 IU/mL.

QFT-Plus Analysis Software was used for qualitative interpretation and quality-control assessment. Laboratory personnel were blinded to the final diagnostic classification. Quantitative TB1−Nil and TB2−Nil responses were retained for secondary analyses.

To assess the incremental contribution of TB2, a TB1-only classification was reconstructed by applying the manufacturer’s antigen-positivity criteria to TB1 alone. This classification was compared with the complete QFT-Plus algorithm, in which either antigen tube could determine positivity. The reconstructed TB1-only classification was not considered analytically equivalent to QFT-GIT.

### Statistical analysis

Categorical variables were summarised as counts and percentages and continuous variables as medians and interquartile ranges. Participant characteristics were compared using Pearson’s chi-squared test for categorical variables and the Wilcoxon rank-sum test for continuous variables. Diagnostic accuracy was evaluated among participants with determinate QFT-Plus results. Sensitivity, specificity, predictive values, accuracy, likelihood ratios, and observed TB prevalence were estimated with 95% confidence intervals (CIs). Wilson intervals were used for proportions and logarithmic intervals for likelihood ratios.

Diagnostic performance was estimated overall and by HIV status, country, and combined country–HIV strata. Participants with unknown HIV status contributed to overall and country estimates but not to HIV-stratified analyses. Sensitivity and specificity were compared between independent groups using two-sample score tests for proportions. The diagnostic information provided by a negative result was expressed primarily using the negative likelihood ratio (LR−). Post-test odds were calculated by multiplying pretest odds by the LR− and then converted to probabilities. Negative predictive values were modelled across hypothetical active-TB prevalences of 0%–40%, holding sensitivity and specificity fixed at the values observed overall and separately among participants with and without HIV. These exploratory scenarios were not interpreted as clinical decision thresholds.

The reconstructed TB1-only classification and complete QFT-Plus algorithm were compared in the same participants with determinate QFT-Plus results. Because the classifications were structurally nested, the contribution of TB2 was described using absolute changes in sensitivity and specificity and the numbers reclassified. CIs for these changes were derived from Wilson intervals for the proportions reclassified among participants with and without active TB, respectively. Paired TB1−Nil and TB2−Nil responses were compared using the Wilcoxon signed-rank test, and comparisons between independent groups used the Wilcoxon rank-sum test.

Indeterminate results were evaluated in the full tested population. Exploratory logistic regression assessed associations with indeterminate results, including country, HIV status, and active TB as covariates. A clinical complete-case model additionally included age and sex. Because of the limited number of events and available measurements, CD4 count, expressed per 100 cells/mm^3^, was evaluated separately among participants with HIV.

No formal sample-size calculation was performed; the sample comprised eligible participants enrolled during the recruitment period. Statistical tests were two-sided, with P<0.05 considered statistically significant. Secondary and subgroup analyses were interpreted as exploratory. Analyses were conducted using R version 4.5.1 (R Foundation for Statistical Computing, Vienna, Austria). Reporting followed the STARD 2015 recommendations [13].

## RESULTS

### Study flow and data completeness

Of 798 adults enrolled in the parent ImmiPrint-TB study, 75 underwent neither QFT-Plus nor microbiological testing and had no diagnosis of active TB; they were excluded from the present analysis. The remaining 723 participants had a QFT-Plus result and a valid final diagnostic classification. Among these, 48 (6.6%) had indeterminate results, leaving 675 participants with determinate results for the primary diagnostic-accuracy analysis: 147 with microbiologically confirmed active TB and 528 without active TB (**Figure 1**).

**Figure 1.**
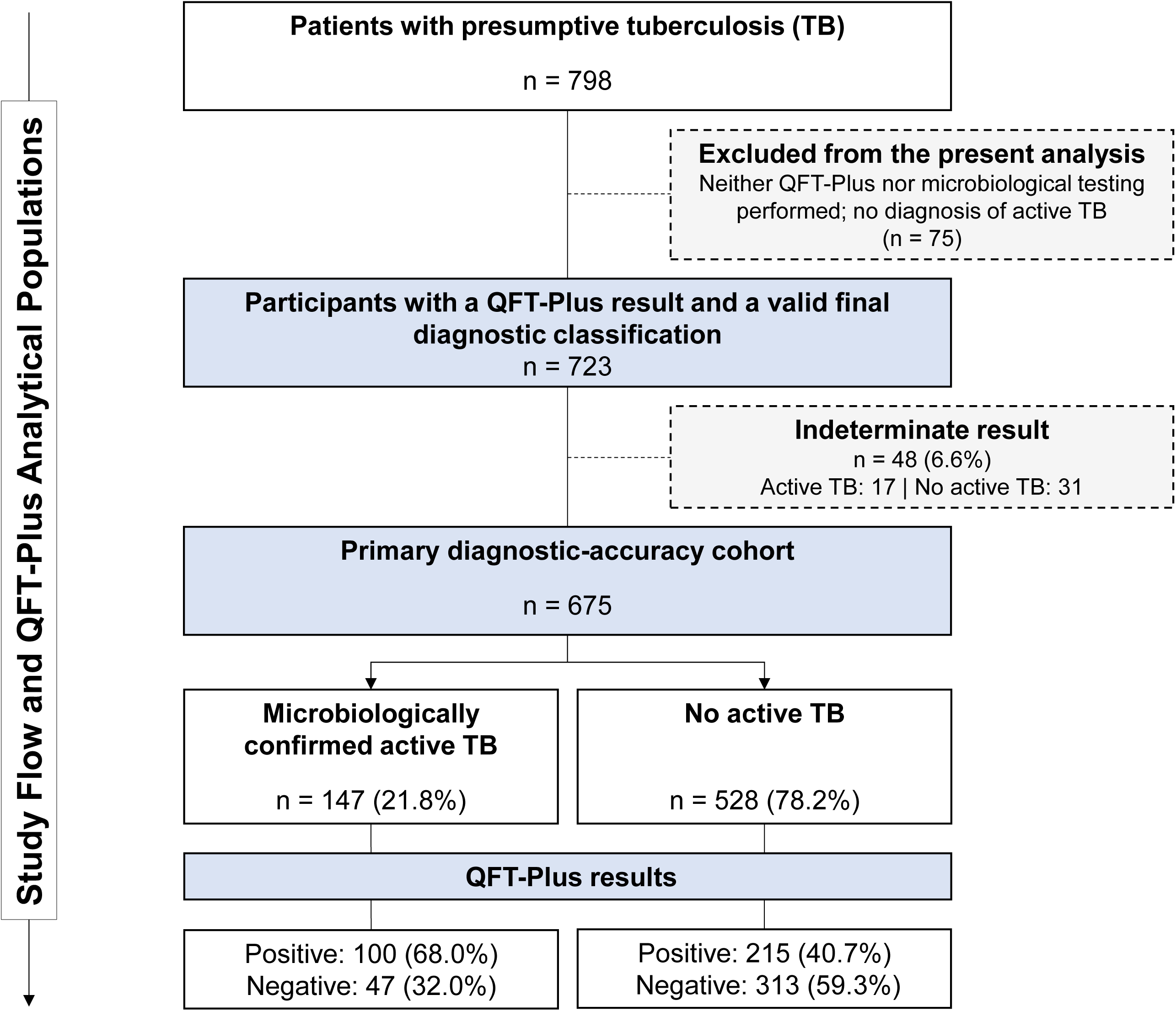
Study flow and QFT-Plus analytical populations. Of 798 adults enrolled in the parent ImmiPrint-TB study, 75 underwent neither QFT-Plus nor microbiological testing and had no diagnosis of active TB; these participants were excluded from the present analysis. The remaining 723 participants had a QFT-Plus result and a valid final diagnostic classification. Forty-eight participants with indeterminate QFT-Plus results were excluded from the primary diagnostic-accuracy analysis but retained for the analysis of indeterminate results. The primary diagnostic-accuracy population comprised 675 participants with determinate results, including 147 with microbiologically confirmed active tuberculosis and 528 without active tuberculosis. TB: Tuberculosis; QFT-Plus: QuantiFERON-TB Gold Plus.

Complete demographic and clinical data were available for 603/675 participants (89.3%), including 385/388 (99.2%) in Brazil and 218/287 (75.9%) in South Africa. HIV status was available for 599/675 participants (88.7%) (**Supplementary Table S1**).

### Participant characteristics

Of the 675 participants, 388 (57.5%) were recruited in Brazil and 287 (42.5%) in South Africa. Among the 603 participants with demographic and clinical data, the median age was 39 years (IQR 30–51), and 373 (61.9%) were male. The most frequent symptoms were cough (530/603, 87.9%), weight loss (387/603, 64.2%), night sweats (299/603, 49.6%), fatigue (264/603, 43.8%), and fever (258/603, 42.8%). Participants with active TB were younger, were more frequently male, and reported night sweats, fatigue, fever, and weight loss more frequently than those without active TB (all P≤0.010). The frequencies of cough and dyspnoea did not differ significantly between groups (**Table 1**).

**Table 1.**
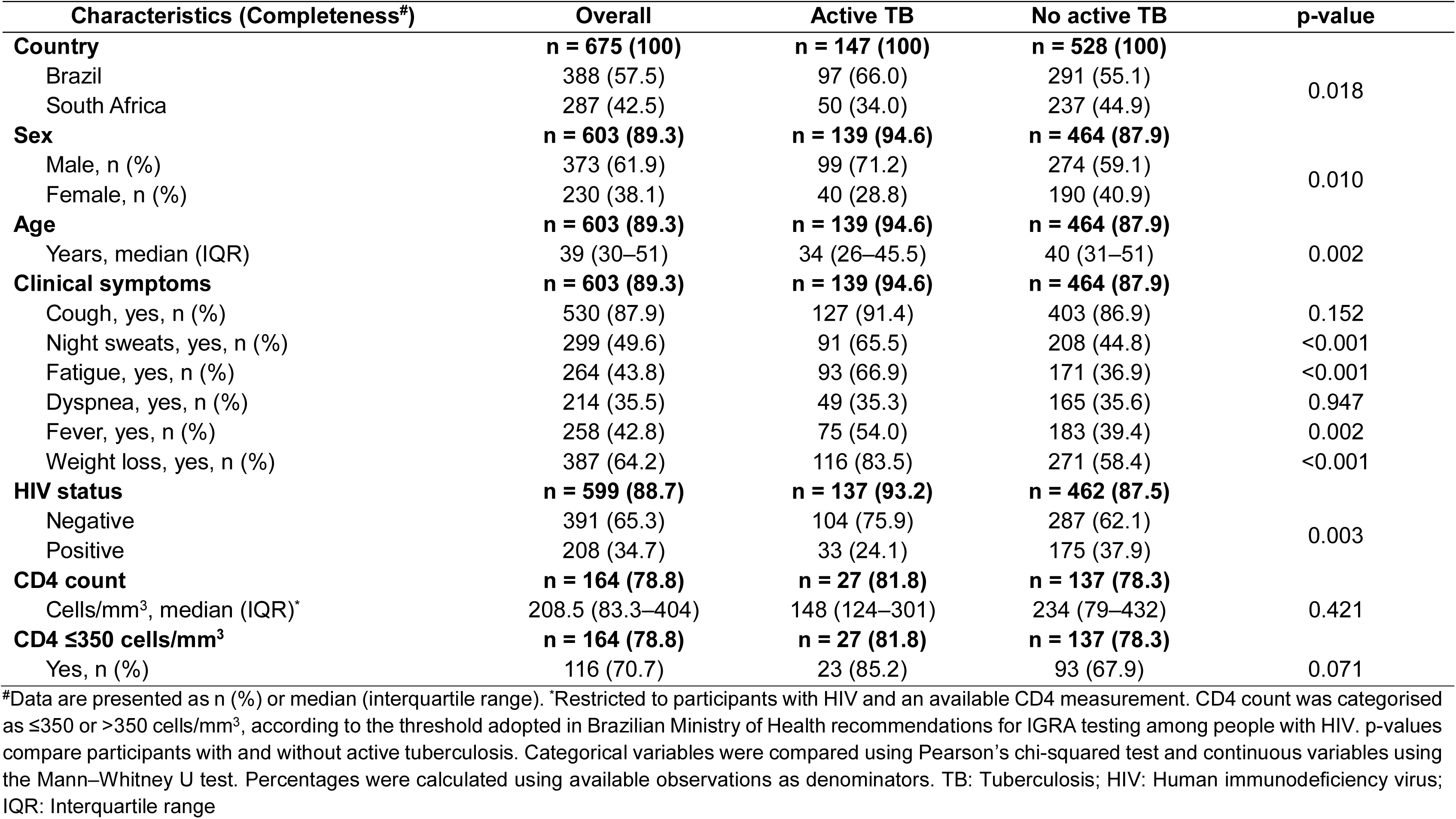
Demographic and clinical characteristics of the primary diagnostic-accuracy cohort according to active tuberculosis status.

| Characteristics (Completeness#) | Overall | Active TB | No active TB | p-value |
| --- | --- | --- | --- | --- |
| **Country** | **n = 675 (100)** | **n = 147 (100)** | **n = 528 (100)** |  |
| Brazil | 388 (57.5) | 97 (66.0) | 291 (55.1) | 0.018 |
| South Africa | 287 (42.5) | 50 (34.0) | 237 (44.9) |  |
| **Sex** | **n = 603 (89.3)** | **n = 139 (94.6)** | **n = 464 (87.9)** |  |
| Male, n (%) | 373 (61.9) | 99 (71.2) | 274 (59.1) | 0.010 |
| Female, n (%) | 230 (38.1) | 40 (28.8) | 190 (40.9) |  |
| **Age** | **n = 603 (89.3)** | **n = 139 (94.6)** | **n = 464 (87.9)** |  |
| Years, median (IQR) | 39 (30–51) | 34 (26–45.5) | 40 (31–51) | 0.002 |
| **Clinical symptoms** | **n = 603 (89.3)** | **n = 139 (94.6)** | **n = 464 (87.9)** |  |
| Cough, yes, n (%) | 530 (87.9) | 127 (91.4) | 403 (86.9) | 0.152 |
| Night sweats, yes, n (%) | 299 (49.6) | 91 (65.5) | 208 (44.8) | <0.001 |
| Fatigue, yes, n (%) | 264 (43.8) | 93 (66.9) | 171 (36.9) | <0.001 |
| Dyspnea, yes, n (%) | 214 (35.5) | 49 (35.3) | 165 (35.6) | 0.947 |
| Fever, yes, n (%) | 258 (42.8) | 75 (54.0) | 183 (39.4) | 0.002 |
| Weight loss, yes, n (%) | 387 (64.2) | 116 (83.5) | 271 (58.4) | <0.001 |
| **HIV status** | **n = 599 (88.7)** | **n = 137 (93.2)** | **n = 462 (87.5)** |  |
| Negative | 391 (65.3) | 104 (75.9) | 287 (62.1) | 0.003 |
| Positive | 208 (34.7) | 33 (24.1) | 175 (37.9) |  |
| **CD4 count** | **n = 164 (78.8)** | **n = 27 (81.8)** | **n = 137 (78.3)** |  |
| Cells/mm<sup>3</sup>, median (IQR)* | 208.5 (83.3–404) | 148 (124–301) | 234 (79–432) | 0.421 |
| **CD4 ≤350 cells/mm<sup>3</sup>** | **n = 164 (78.8)** | **n = 27 (81.8)** | **n = 137 (78.3)** |  |
| Yes, n (%) | 116 (70.7) | 23 (85.2) | 93 (67.9) | 0.071 |

Among 599 participants with known HIV status, 208 (34.7%) were living with HIV. CD4 counts were available for 164 participants with HIV, with a median of 208.5 cells/mm³ (IQR 83.3–404); 116/164 (70.7%) had CD4 ≤350 cells/mm³. In exploratory HIV-stratified analyses, participants with HIV and active TB reported night sweats and weight loss more frequently than those with HIV but without active TB. Among participants without HIV, active TB was additionally associated with younger age, male sex, fatigue, and fever (**Supplementary Table S2**). Country-stratified comparisons of participant characteristics by TB status are presented in **Supplementary Table S3**.

### Diagnostic information provided by QFT-Plus

Among 675 participants with determinate results, the observed prevalence of active TB was 21.8% (147/675; 95% CI, 18.8–25.0). QFT-Plus sensitivity was 68.0% (100/147; 95% CI, 60.1–75.0), with negative results in 47 participants with active TB. Specificity was 59.3% (313/528; 95% CI, 55.0–63.4), and the negative likelihood ratio (LR−) was 0.54 (95% CI, 0.42–0.69) (**Table 2**). Based on this point estimate, a negative result reduced disease odds by 46% but, at the observed prevalence, left an estimated post-test probability of active TB of 13.1%.

**Table 2.** Diagnostic accuracy of QFT-Plus for active tuberculosis in the primary cohort.

| Population<sup>#</sup> | TP/FN/FP/TN | TB Prevalence, %<br>(95% CI) | Sensitivity, %<br>(95% CI) | Specificity, %<br>(95% CI) | PPV, %<br>(95% CI) | NPV, %<br>(95% CI) | LR+<br>(95% CI) | LR-<br>(95% CI) |
| --- | --- | --- | --- | --- | --- | --- | --- | --- |
| Overall (n = 675) | 100/47/215/313 | 21.8<br>(18.8–25.0) | 68.0<br>(60.1–75.0) | 59.3<br>(55.0–63.4) | 31.7<br>(26.9–37.1) | 86.9<br>(83.1–90.0) | 1.67<br>(1.44–1.94) | 0.54<br>(0.42–0.69) |
| HIV-positive (n = 208) | 23/10/48/127 | 15.9<br>(11.5–21.4) | 69.7<br>(52.7–82.6) | 72.6<br>(65.5–78.6) | 32.4<br>(22.7–43.9) | 92.7<br>(87.1–96.0) | 2.54<br>(1.83–3.53) | 0.42<br>(0.25–0.71) |
| HIV-negative (n = 391) | 69/35/145/142 | 26.6<br>(22.5–31.2) | 66.3<br>(56.8–74.7) | 49.5<br>(43.7–55.2) | 32.2<br>(26.3–38.8) | 80.2<br>(73.7–85.4) | 1.31<br>(1.10–1.57) | 0.68<br>(0.51–0.91) |
| Brazil (n = 388) | 67/30/85/206 | 25.0<br>(21.0–29.5) | 69.1<br>(59.3–77.4) | 70.8<br>(65.3–75.7) | 44.1<br>(36.4–52.0) | 87.3<br>(82.4–90.9) | 2.36<br>(1.89–2.96) | 0.44<br>(0.32–0.59) |
| Brazil, HIV-positive (n = 122) | 11/3/23/85 | 11.5<br>(7.0–18.3) | 78.6<br>(52.4–92.4) | 78.7<br>(70.1–85.4) | 32.4<br>(19.1–49.2) | 96.6<br>(90.5–98.8) | 3.69<br>(2.34–5.81) | 0.27<br>(0.10–0.75) |
| Brazil, HIV-negative (n = 190) | 48/25/40/77 | 38.4<br>(31.8–45.5) | 65.8<br>(54.3–75.6) | 65.8<br>(56.8–73.8) | 54.5<br>(44.2–64.5) | 75.5<br>(66.3–82.8) | 1.92<br>(1.42–2.60) | 0.52<br>(0.37–0.73) |
| South Africa (n = 287) | 33/17/130/107 | 17.4<br>(13.5–22.2) | 66.0<br>(52.2–77.6) | 45.1<br>(38.9–51.5) | 20.2<br>(14.8–27.1) | 86.3<br>(79.1–91.3) | 1.20<br>(0.96–1.51) | 0.75<br>(0.50–1.14) |
| South Africa, HIV-positive<br>(n = 86) | 12/7/25/42 | 22.1<br>(14.6–31.9) | 63.2<br>(41.0–80.9) | 62.7<br>(50.7–73.3) | 32.4<br>(19.6–48.5) | 85.7<br>(73.3–92.9) | 1.69<br>(1.07–2.69) | 0.59<br>(0.32–1.09) |
| South Africa, HIV-negative<br>(n = 201) | 21/10/105/65 | 15.4<br>(11.1–21.1) | 67.7<br>(50.1–81.4) | 38.2<br>(31.3–45.7) | 16.7<br>(11.2–24.1) | 86.7<br>(77.2–92.6) | 1.10<br>(0.84–1.44) | 0.84<br>(0.49–1.45) |

<sup>#</sup>Indeterminate QFT-Plus results were excluded from the diagnostic-accuracy analysis. Confidence intervals for proportions were calculated using the Wilson method, and confidence intervals for likelihood ratios were calculated using the logarithmic method. HIV status was unknown or not performed for 76 participants, who were included in the overall and country analyses but excluded from HIV-stratified estimates. TP: True positive; FN: False negative; FP: False positive; TN: True negative; TB: Tuberculosis; HIV: Human immunodeficiency virus; PPV: Positive predictive value; NPV: Negative predictive value; LR+: Positive likelihood ratio; LR-: Negative likelihood ratio.

Sensitivity did not differ significantly between participants with and without HIV (69.7% versus 66.3%; P=0.72). Specificity was descriptively higher among participants with HIV (72.6% versus 49.5%), and LR− was 0.42 (95% CI, 0.25–0.71) and 0.68 (95% CI, 0.51–0.91), respectively. The higher negative predictive value (NPV) among participants with HIV (92.7% versus 80.2%) occurred alongside lower active-TB prevalence (15.9% versus 26.6%) and higher specificity; nevertheless, 10/33 participants with HIV and active TB had negative QFT-Plus results. Sensitivity did not differ significantly between Brazil and South Africa (69.1% versus 66.0%; P=0.71), whereas specificity was lower in South Africa (45.1% versus 70.8%; P<0.001). The LR− in South Africa was 0.75 (95% CI, 0.50–1.14) (**Table 2**).

Across the four country–HIV strata, LR− ranged from 0.27 (95% CI, 0.10–0.75) among participants with HIV in Brazil to 0.84 (95% CI, 0.49–1.45) among participants without HIV in South Africa. The LR− confidence intervals included 1 in both South African strata and were therefore compatible with no change in disease odds after a negative result (**Table 2**; **Figure 2A**). Specificity was descriptively higher among participants with HIV in both countries. NPV ranged from 75.5% among participants without HIV in Brazil, where active-TB prevalence was 38.4%, to 96.6% among participants with HIV in Brazil, where prevalence was 11.5%. With sensitivity and specificity held fixed at the overall estimates, modelled NPV reached 90% only at active-TB prevalences below approximately 17% (**Supplementary Figure S1**).

**Figure 2.**
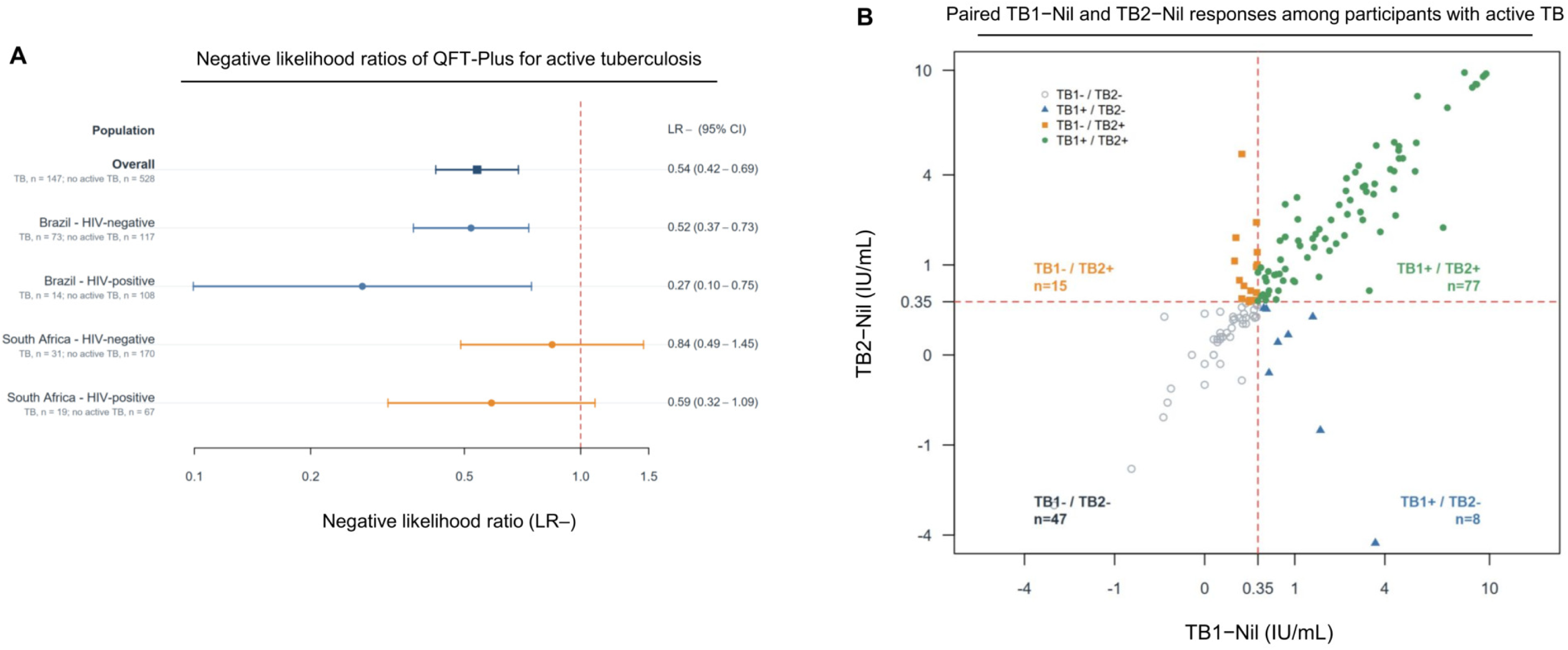
Negative likelihood ratios and paired TB1−Nil and TB2−Nil responses according to active tuberculosis status. (A) Negative likelihood ratios for QFT-Plus in the overall population and in strata defined by country and HIV status. Points represent estimates and horizontal lines represent 95% confidence intervals calculated using the log-likelihood-ratio approximation. The vertical dashed line indicates LR− = 1. (B) Paired TB1−Nil and TB2−Nil interferon-gamma concentrations among participants with microbiologically confirmed active tuberculosis. Lines connect paired measurements from the same participant; symbols indicate TB1/TB2 positivity patterns according to the manufacturer’s criteria. QFT-Plus: QuantiFERON-TB Gold Plus; LR−: Negative likelihood ratio; CI: Confidence interval; TB: Tuberculosis.

### Indeterminate QFT-Plus results

All 48 indeterminate results (6.6%; 95% CI, 5.0–8.7) were attributable to insufficient mitogen responses. Indeterminate results were more frequent among participants with HIV than among those without HIV (26/234, 11.1%, versus 20/411, 4.9%) and among participants with active TB than among those without active TB (17/164, 10.4%, versus 31/559, 5.5%) (**Supplementary Table S4**).

In the exploratory model including country, HIV status, and active TB, both HIV infection (adjusted odds ratio [aOR], 2.78; 95% CI, 1.49–5.15; P=0.001) and active TB (aOR, 2.28; 95% CI, 1.21–4.31; P=0.011) were associated with higher odds of an indeterminate result after mutual adjustment. Country was not significantly associated with indeterminate results (South Africa versus Brazil: aOR, 1.45; 95% CI, 0.79–2.68; P=0.23) (**Supplementary Table S5**).

### Incremental contribution of the TB2 antigen tube

Compared with the reconstructed TB1-only classification, the complete QFT-Plus algorithm increased sensitivity from 57.8% (85/147; 95% CI, 49.7–65.5) to 68.0% (100/147; 95% CI, 60.1–75.0), an absolute gain of 10.2 percentage points (95% CI, 6.3–16.2) corresponding to 15 additional participants with active TB testing positive. Specificity decreased from 62.7% (331/528; 95% CI, 58.5–66.7) to 59.3% (313/528; 95% CI, 55.0–63.4), an absolute reduction of 3.4 percentage points (95% CI, 2.2–5.3) corresponding to 18 additional positive results among participants without active TB (**Table 3**; **Figure 2B; Supplementary Figure S2**).

**Table 3.**
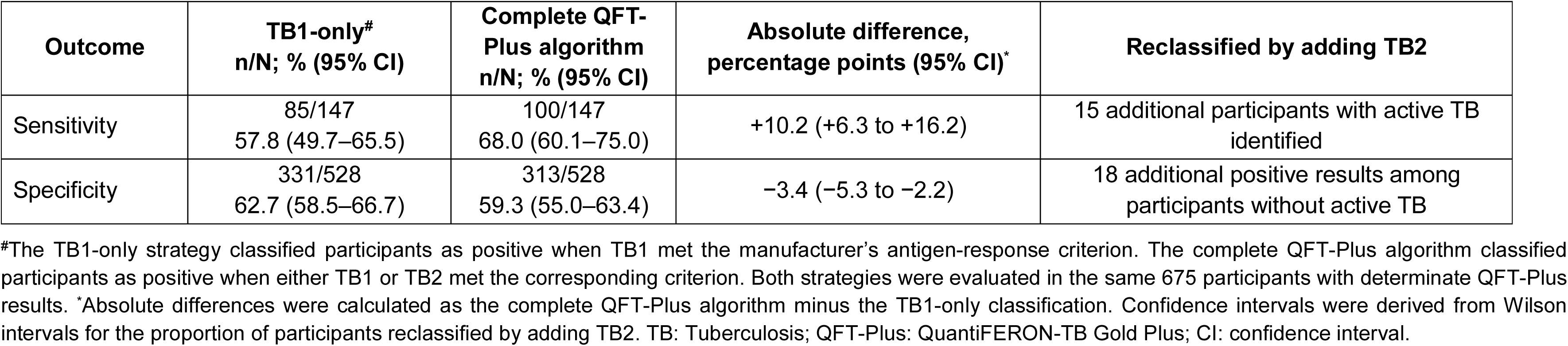
Incremental contribution of TB2 to QFT-Plus classification.

### Quantitative TB1 and TB2 responses and positivity patterns

Among participants in the primary diagnostic-accuracy analysis, median TB1−Nil and TB2−Nil concentrations were 0.20 and 0.19 IU/mL, respectively, with no statistically significant paired difference (P=0.81). No significant paired differences were observed within strata defined by active-TB or HIV status. Country-specific differences were small and occurred in opposite directions: median TB2−Nil was higher than TB1−Nil in Brazil (0.06 versus 0.05 IU/mL; P<0.001) but lower in South Africa (0.44 versus 0.51 IU/mL; P<0.001). Similar patterns were observed in the full tested population (**Supplementary Figure S3; Supplementary Tables S6 and S7**).

Among the 675 participants with determinate results, 360 were negative in both antigen tubes, 33 were positive only in TB1, 33 were positive only in TB2, and 249 were positive in both. Among participants with active TB, the corresponding counts were 47, 8, 15, and 77, respectively; among those without active TB, they were 313, 25, 18, and 172 (**Figure 2B; Supplementary Figure S4**).

## DISCUSSION

In this prospective diagnostic-accuracy study of adults investigated for TB in Brazil and South Africa, a negative QFT-Plus result reduced disease odds but left substantial residual uncertainty. At the observed prevalence, the post-test probability of active TB remained 13.1%, and approximately one-third of participants with microbiologically confirmed TB and determinate results tested negative. The information provided by a negative result varied across country–HIV strata, with confidence intervals compatible with no change in disease odds in both South African strata. These findings highlight the importance of interpreting negative results in the context of the population tested. Adding TB2 improved case detection relative to the reconstructed TB1-only classification but did not overcome the limited sensitivity of the complete algorithm. Indeterminate results were more frequent among participants with HIV and among those with active TB, further limiting the proportion receiving an interpretable result in these groups.

The overall sensitivity of 68.0% was consistent with the suboptimal IGRA performance previously reported among patients evaluated for active TB in high-burden and high-HIV-prevalence settings [3,8,9]. QFT sensitivity also varies according to immune status. Studies of culture-confirmed TB have reported lower sensitivity among immunocompromised participants, including estimates around 70% [14,15]. These findings support the interpretation that impaired cellular immunity may reduce antigen-specific responses and that a negative QFT-Plus result cannot reliably exclude active TB. This is consistent with WHO guidance, which positions IGRAs as tests for TB infection rather than for diagnosing or excluding TB disease [2].

The overall NPV of 86.9% should not be interpreted as an intrinsic rule-out property of QFT-Plus. The higher NPV among participants with HIV occurred alongside lower active-TB prevalence and higher specificity, without a statistically significant difference in sensitivity. Nevertheless, 10/33 participants with HIV and microbiologically confirmed active TB had negative results. When sensitivity and specificity were held fixed at the overall estimates, modelled NPV reached 90% only at active-TB prevalences below approximately 17%. This projection illustrates the dependence of NPV on prevalence; it does not establish a safe threshold for excluding disease. Neither the overall nor HIV-specific NPV therefore supports ending diagnostic investigation on the basis of a negative QFT-Plus result alone.

The incremental contribution of TB2 requires a more nuanced interpretation. Inclusion of TB2 increased sensitivity from 57.8% to 68.0%, identifying 15 additional TB cases, but generated 18 additional positive results among participants without active TB and reduced specificity from 62.7% to 59.3%. Systematic reviews comparing QFT-Plus with QFT-GIT have similarly found high agreement and, at most, small differences in sensitivity [7,16]. However, the reconstructed TB1-only classification is not analytically equivalent to QFT-GIT and should not be interpreted as a direct comparison between assays. Rather, it quantifies the within-assay contribution of TB2 to the complete QFT-Plus algorithm.

Paired TB1−Nil and TB2−Nil responses did not differ overall or within active-TB and HIV strata. Small country-specific differences occurred in opposite directions and may reflect differences in study populations, pre-analytical conditions, or laboratory procedures. Although TB2 contains additional peptides intended to elicit CD8^+^ and CD4^+^ T-cell responses, neither antigen tube is cell specific and TB2−TB1 is not a direct measure of the *M. tuberculosis*-specific CD8^+^ response [5]. Experimental evidence that both cell populations contribute to TB2 responses further limits mechanistic interpretation [4]. These quantitative findings should therefore be considered exploratory.

The low specificity for active TB is consistent with QFT-Plus detecting immunological sensitisation to *M. tuberculosis* without distinguishing infection from active disease [2, 4]. This limits the interpretation of positive results in high-burden settings and reinforces the need to quantify the information provided by a negative result. Specificity was lower in South Africa than in Brazil, whereas sensitivity did not differ significantly between countries. Differences in background prevalence of TB infection, participant selection, clinical case mix, referral patterns, and site-level procedures could have contributed to this variation, but their individual contributions could not be determined. Specificity was descriptively higher among participants with HIV in both countries. Reduced antigen-specific responsiveness among participants with HIV without active TB is one possible explanation, but this mechanism was not directly investigated.

Indeterminate results further limited the diagnostic value of QFT-Plus. In the exploratory regression model, HIV and active TB were associated with higher odds of an indeterminate result after adjustment for each other and country. These associations highlight that failure to obtain an interpretable result was more frequent in clinically relevant groups, although the analysis does not establish the underlying mechanisms. Previous studies have reported reduced sensitivity and more frequent indeterminate results among immunocompromised patients with active TB and have linked indeterminate QFT-Plus results to lower lymphocyte, CD4^+^, and CD8^+^ T-cell counts [17, 18]. Indeterminate results should be explicitly considered when assessing diagnostic performance and should not be interpreted as evidence against active TB. When disease remains suspected, clinical, radiological, and microbiological investigation should continue.

Study strengths included prospective enrolment of adults undergoing investigation for active TB, separation of the index test from clinical decision-making, blinded laboratory interpretation, and microbiological confirmation of active TB. Participants classified as not having active TB required an alternative diagnosis based on medical assessment, laboratory findings, and clinical follow-up. Inclusion of sites in Brazil and South Africa enabled evaluation across populations with different referral profiles and healthcare settings.

Several limitations should be considered. Recruitment relied on an operational convenience sample, which may limit representativeness, and lower clinical-data completeness in South Africa constrained adjusted and subgroup analyses. Restricting the active-TB group to microbiologically confirmed cases prioritised reference-standard specificity but limited generalisability to patients with clinically diagnosed TB without microbiological confirmation, including some with extrapulmonary or paucibacillary disease. Primary accuracy estimates were conditional on a determinate QFT-Plus result and may therefore overstate clinical utility in the full tested population, although indeterminate results were analysed separately. Some country–HIV strata included few TB cases, resulting in imprecise estimates. The study did not evaluate clinical outcomes, cost-effectiveness, or QFT-Plus-guided decision-making. The incremental contribution of TB2 and the quantitative response patterns require confirmation in independent populations.

These findings have practical implications for TB services, including in Brazil, where IGRAs are recommended primarily for the investigation of TB infection rather than active disease. In patients with symptoms or other clinical features suggestive of active TB, negative or indeterminate QFT-Plus results should not delay radiological assessment, rapid molecular testing, culture, or collection of additional specimens when indicated. Interpretive statements in laboratory reports and clinical protocols may help prevent inappropriate diagnostic closure after negative or indeterminate results, particularly among people with HIV.

In conclusion, a negative QFT-Plus result provided insufficient information to exclude active TB among adults investigated in Brazil and South Africa. The higher NPV among participants with HIV occurred alongside lower disease prevalence and higher specificity, without evidence of greater sensitivity. Adding TB2 increased case detection but did not overcome the limited sensitivity of the complete algorithm. Negative or indeterminate QFT-Plus results alone should not terminate diagnostic evaluation when active TB remains suspected.

## CONFLICT OF INTEREST

The authors have declared that no competing interests exist.

## FUNDING

The study was supported by the European Union, the Coordenação de Aperfeiçoamento de Pessoal de Nível Superior (CAPES), Conselho Nacional de Desenvolvimento Científico e Tecnológico (CNPq) (CNPq/DECIT/MS #31/2024 – Process #442411/2024-3, CNPq/PV #49/2024 - Process #351706/2025-9 and CNPq/MCTI Universal #44/2024 - Process: 408941/2025-1), Fundação de Amparo à Pesquisa do Estado do Amazonas (FAPEAM) (PECTI-AM/SAÚDE Program - #004/2020, INOVATEC+ Program - #015/2024, FRONTEIRAS DO CONHECIMENTO/FAPEAM Program #015/2025 and POSGRAD Program #015/2026), South African Medical Research Council (SA MRC) (RFA-EMU-02-2017), European and Developing Countries Clinical Trials Partnership (EDCTP) (TMA-2015SF-1043, TMA-1051-TESAII and TMA-CDF2015), United Kingdom Medical Research Council (UK MRC) (MR/S03563X/1) and the Wellcome Trust (MR/S027777/1). AGC, AT, VSS and MC-S are research fellows of CNPq. AGC is also a research fellow supported by CNPq (Visiting Researcher Program #049-2024). The funders had no role in the study design, data collection, data analysis, interpretation of the findings, preparation of the manuscript or decision to submit the article for publication.

## ACKNOWLEDGEMENTS

We thank all participants and healthcare professionals involved in participant care and study procedures at Fundação de Medicina Tropical Dr Heitor Vieira Dourado (FMT-HVD), Fundação de Pneumologia Sanitária Cardoso Fontes (FCF), and the participating primary healthcare centres in Cape Town.

## DATA AVAILABILITY STATEMENT

The de-identified participant-level data underlying this study are not publicly available because of ethical and confidentiality restrictions. Requests for access should be directed to the Research Ethics Committee of Fundação de Medicina Tropical Doutor Heitor Vieira Dourado (CEP/FMT-HVD; telephone: +55 92 2127-3572). The corresponding author is available to facilitate enquiries. Access will be considered subject to review of the proposed use, approval by the relevant institutional ethics committee, and compliance with applicable data protection requirements.

## AUTHORS’ CONTRIBUTIONS

*Conceptualization:* Anete Trajman, Keertan Dheda and Marcelo Cordeiro-Santos.

*Methodology:* Allyson Guimarães Costa, Maria Cecília Borges-Cabral, Alexandra Brito Souza and Renata Spener-Gomes.

*Formal analysis:* Allyson Guimarães Costa, Maria Cecília Borges-Cabral, Vanderson Souza Sampaio and Daniel Barros de Castro.

*Investigation:* Allyson Guimarães Costa, Maria Cecília Borges-Cabral, Aliasgar Esmail, Brenda Karoline Souza Carvalho, Bruna Pires Loiola, Alexandra Brito Souza, Renata Spener-Gomes and Jaquelane Silva Jesus.

*Resources:* Anete Trajman, Keertan Dheda and Marcelo Cordeiro-Santos.

*Data curation:* Allyson Guimarães Costa, Maria Cecília Borges-Cabral, Vanderson Souza Sampaio and Daniel Barros de Castro.

*Writing – Original Draft:* Allyson Guimarães Costa and Maria Cecília Borges-Cabral.

*Writing – Review & Editing:* Anete Trajman, Aliasgar Esmail, Keertan Dheda and Marcelo Cordeiro-Santos.

*Visualization:* Allyson Guimarães Costa and Maria Cecília Borges-Cabral.

*Supervision:* Anete Trajman, Keertan Dheda and Marcelo Cordeiro-Santos.

*Project administration:* Allyson Guimarães Costa, Alexandra Brito Souza and Renata Spener-Gomes.

*Funding acquisition:* Anete Trajman, Aliasgar Esmail, Keertan Dheda and Marcelo Cordeiro-Santos.

## Notes

### Competing Interest Statement

The authors have declared no competing interest.

### Author Declarations

The Brazilian component was approved by the Research Ethics Committee of Fundacao de Medicina Tropical Doutor Heitor Vieira Dourado (approval no. 2,525,182; CAAE 80643917.4.0000.5260), and the South African component by the University of Cape Town Human Research Ethics Committee (HREC reference 290/2020). All procedures complied with applicable national regulations and the Declaration of Helsinki. All participants provided written informed consent before enrolment.

